# Comprehensive Usability Evaluation and Performance Overview of Pathpoint® Outcomes Software

**DOI:** 10.1101/2024.05.11.24307045

**Authors:** Balamurugan Subramaniyan, Atlas Naqvi, Muna Mohamud, Piyush Mahapatra

## Abstract

This research evaluates the usability of Pathpoint® Outcomes Software, aligning with IEC 62366-1:2015 standards to ensure rigour and accuracy. The study involved diverse user groups like Healthcare Professionals and the General Population to provide varied perspectives. The study focused on the software’s usability, safety, and effectiveness, which are crucial to user experience, satisfaction, and performance. Any potential risks to users or patient data were identified to ensure safety standards. The software’s effectiveness was assessed, considering its accuracy, reliability, and efficiency. The study used both quantitative metrics and qualitative feedback for a balanced evaluation. It also aimed to identify and mitigate potential use errors, enhancing the software’s usability, safety, and effectiveness.

## 1 Introduction

In the ever-evolving and dynamic landscape of medical device development, the importance of prioritising usability has become a critical imperative. This is a fact that has been emphasised by the International Electrotechnical Commission’s (IEC) standard 62366. This standard is a crucial guide that directs the integration of usability engineering processes throughout the various stages of medical device design and development [ISO, 2015; Wiklund and Kendler, 2018; IEC, 2016; Karshmer and Tenenberg, 2016].

The importance of usability studies in the field of medical device development is well established, as evidenced by research conducted by Bevan and Macleod in 1994. Their work underscores the importance of considering productivity, effectiveness, and user satisfaction for the evaluation of usability. They also delved into exploratory methodologies for evaluating usability, providing a comprehensive understanding of the subject. The ESPRIT MUSiC project, which contributed tools for measuring usability in both lab and field environments, is another significant contribution to the field [Virzi, 1992].

Foundational works by Virzi in 1992, Nielsen in 1993, and Lewis in 1995 have significantly contributed to the field of usability studies. They have provided valuable insights into optimising the test phase of usability studies and validated tools for measuring user satisfaction. These works have been instrumental in shaping the direction of usability.

The introduction of the System Usability Scale (SUS) by Brooke in 1996 has been a significant milestone in the field of usability studies. This tool is quick and easy to use, making it a valuable asset for researchers and developers alike [Brooke, 1996]. Dumas and Redish’s comprehensive guide to usability testing, published in 1999, is another important resource that provides a detailed guide to conducting usability tests [Dumas and Redish, 1999]. Shneiderman’s influential book on designing user interfaces, published in 1998, is a treasure trove of knowledge and methodologies in the field [Shneiderman, 1998].

Building upon this solid foundation, the work by Kushniruk and Patel in 2004 focuses on cognitive and usability engineering methods in the evaluation of clinical information systems. Their work provides a comprehensive understanding of the subject and offers valuable insights into the field [Kushniruk and Patel, 2004]. The study by Marcilly, Ammenwerth, and Roehrer in 2006 assesses the usability of user interfaces for clinical information systems and explores effective evaluation frameworks [Marcilly et al., 2006]. Rubin and Chisnell’s Handbook, published in 2008, offers practical methodologies for conducting effective usability tests [Rubin and Chisnell, 2008].

The intersection of usability studies and medical devices is a contemporary research focus. This is demonstrated by the work of Patel et al., who applied usability testing within the context of IEC 62366 [Patel et al., 2017]. Hignett et al.’s emphasis on usability engineering in medical device development is another significant contribution to the field [Hignett et al., 2018]. The exploration of IEC 62366 integration by Nielsen et al. and the review of usability evaluation methodologies and IEC 62366 compliance by Smith et al. highlight ongoing efforts to align usability practices with regulatory standards [Nielsen et al., 2019; Smith et al., 2020].

In this context, our research aims to contribute to this body of knowledge by presenting a comprehensive usability evaluation and performance overview of the Pathpoint® Outcomes Software. This study is grounded in the principles of IEC 62366 and is informed by established usability methodologies. This study aims to enhance the overall usability and user experience of medical technologies. We aim to improve patient outcomes and user satisfaction by aligning with regulatory standards. This research is a significant step towards achieving these goals and contributes to the ongoing efforts to improve the usability of medical devices.

## 2 Methods

The software’s usability processes and documentation were meticulously mapped to IEC 62366-1:2015 requirements to ensure adherence to established standards. This was a crucial step in the process, as it ensured that the software was developed in line with internationally recognized standards for usability. The IEC 62366-1:2015 standard is a comprehensive guideline that provides a framework for the application of usability engineering to medical devices. It outlines the necessary steps and procedures to ensure that the software is user-friendly, safe, and effective for its intended users.

The meticulous mapping of the software’s usability processes and documentation to the IEC 62366-1:2015 requirements involved a detailed analysis and comparison of the software’s features and functionalities with the standard’s requirements. This process was not only rigorous but also systematic, ensuring that every aspect of the software’s usability was thoroughly evaluated and aligned with the standard.

Key sections, including Information for Safety related to Usability and the Usability Engineering File, were integral to the evaluation process. These sections provided crucial information that was necessary for the comprehensive evaluation of the software’s usability.

The Information for Safety related to Usability section is critical to the IEC 62366-1:2015 standard. It focuses on providing users with necessary information about the software’s safe and effective use. This section includes detailed information about potential hazards, warnings, and precautions that users must be aware of when using the software. It is designed to ensure that users have all the necessary information to use the software safely and effectively.

The Usability Engineering File, on the other hand, is a comprehensive document that contains all the relevant information related to the usability of the software. It includes detailed documentation of the usability engineering processes, methods, and results that have been documented throughout the development lifecycle of the software. This file serves as a comprehensive record of the software’s usability, providing a detailed overview of the software’s usability features and functionalities.

Usability activities followed the risk management procedure, guided by essential documents like the Hazard-Related Use Scenarios list, Usability Evaluation Protocol, and Usability Evaluation Plan. This approach ensured that the usability activities were conducted systematically and controlled, ensuring the overall safety and effectiveness of the software.

The risk management procedure is a systematic approach to identifying, assessing, and mitigating usability risks. It involves a detailed analysis of the software’s features and functionalities, identifying potential usability risks, assessing the severity of these risks, and implementing appropriate measures to mitigate these risks. This approach ensures that potential harm to users is minimized and that the overall safety of the software is ensured.

Several essential documents were utilized to support the risk management process. One such document was the Hazard-Related Use Scenarios list. This list identifies potential hazards that users may encounter while using the software and describes the corresponding use scenarios. By identifying and understanding these hazards, appropriate usability measures can be implemented to mitigate the associated risks.

The Usability Evaluation Protocol was another essential document that was used in the risk management process. This protocol outlines the specific procedures and methods for conducting usability evaluations. It ensures consistency and repeatability in the evaluation process, ensuring that the software’s usability is evaluated in a consistent and reliable manner.

The Usability Evaluation Plan, on the other hand, provides a comprehensive overview of the entire usability evaluation process. This plan includes details about the objectives, scope, methods, participants, and schedule of the evaluation. It serves as a roadmap for conducting the evaluations, ensuring that all necessary aspects of the usability evaluation process are covered.

By following these established standards, conducting usability activities in accordance with a risk management procedure, and utilizing essential documents such as the Hazard-Related Use Scenarios list, Usability Evaluation Protocol, and Usability Evaluation Plan, the software’s usability was thoroughly evaluated and ensured to meet the necessary standards for safety and effectiveness.

This comprehensive approach to usability evaluation ensures that the software is not only user-friendly but also safe and effective for its intended users. It ensures that the software meets the highest standards of usability, providing users with a reliable, safe, and effective tool for their needs. This approach to usability evaluation is not only rigorous but also systematic, ensuring that every aspect of the software’s usability is thoroughly evaluated and aligned with the highest standards of usability.

## 3 User Groups and Preparation

The study that was conducted encompassed two distinct user groups: Healthcare Professionals (HCPs) and the General Population. These two groups were chosen because they represent the primary users of the system or software being evaluated. The Healthcare Professionals group consisted of individuals who work in the healthcare field and use the software or system as part of their daily responsibilities. The general population group, on the other hand, consists of individuals who are not healthcare professionals but may interact with the software or system indirectly, such as patients or their family members.

The participants in the study were encouraged to review the user manual thoroughly. This manual was designed to provide detailed instructions and guidelines on navigating and using the software or system being evaluated. It was created with the intention of being a comprehensive resource for users, providing them with all the information they need to use the software or system effectively. The user manual was designed to be user-friendly and easy to understand, with clear instructions and explanations.

In addition to the user manual, optional training sessions were provided to the participants. These training sessions were designed to provide further guidance and enhance the participants’ understanding of the software or system. The training sessions were conducted by experts who have a deep understanding of the software or system and were able to provide detailed explanations and demonstrations. The participants were given the choice to participate in these training sessions, and it was encouraged but not mandatory.

The tasks assigned to the Healthcare Professionals group primarily focused on data management, communication, and Electronic Health Record (EHR) interaction. These tasks were chosen because they represent the core responsibilities of healthcare professionals when using the software or system. The tasks included entering and retrieving patient data, communicating with colleagues or patients through the software, and effectively using the EHR features. The aim of these tasks was to evaluate the efficiency and ease of use of the software or system in the context of healthcare professionals’ daily responsibilities.

The General Population group was assigned tasks that centred around questionnaire interactions. These tasks were designed to assess how easily individuals from the general population could navigate through questionnaires, provide accurate responses, and interact with the software or system to complete the required tasks. The questionnaires were designed to be user-friendly and easy to understand, with clear instructions and explanations. The aim of these tasks was to evaluate the usability and effectiveness of the software or system for individuals who are not healthcare professionals but may interact with the software or system indirectly.

The study aimed to examine the usability and effectiveness of the user manual and optional training for these two distinct user groups: Healthcare Professionals and the General Population. The goal was to determine if the provided resources adequately supported both user groups in effectively utilising the software or system for their respective needs. The study also sought to gather comprehensive feedback and insights into the usability and effectiveness of the user manual and optional training.

The results of the study would help identify any areas of improvement or potential modifications needed to enhance the user experience and overall usability of the software or system. The feedback and insights gathered from the study would be invaluable in making necessary adjustments and improvements to the software or system. The ultimate goal is to ensure that the software or system is user-friendly and effective for all users, regardless of their background or level of expertise.

By including both Healthcare Professionals and the General Population in the study, the researchers aimed to gather a comprehensive understanding of the usability and effectiveness of the user manual and optional training. This approach ensured that the feedback and insights gathered were representative of the diverse range of users who interact with the software or system. The inclusion of these two distinct user groups in the study was crucial in ensuring that the results were comprehensive and representative of the diverse range of users who interact with the software or system.

In summary, the study aimed to evaluate the usability and effectiveness of the user manual and optional training for two distinct user groups: Healthcare Professionals and the General Population. The participants were encouraged to thoroughly review the user manual and participate in optional training sessions. The tasks assigned to the participants were designed to evaluate the efficiency and ease of use of the software or system in the context of their respective responsibilities. The results of the study would help identify areas of improvement and potential modifications needed to enhance the user experience and overall usability of the software or system.

**Table 1.** User Groups.

| Table 1 |  | User Groups |  |  |
| --- | --- | --- | --- | --- |
| Type of the User | Total Participants | Mean Age | Gender (M/F) | No. of Successful Participants |
| General Population | 10 | 35 | (M-5/F-5) | 10 |
| Healthcare Professionals | 15 | 35 | (M-4/F-11) | 15 |

## 4 Task Selection

In the process of designing the usability study, a significant amount of attention and thought was dedicated to the selection of tasks. The primary aim was to conduct a comprehensive assessment of the Software as a Medical Device (SaMD) and its user interface. The tasks chosen were not random but were meticulously selected to evaluate a wide range of aspects. These aspects included user interactions with the software, the responsiveness of the system to user inputs, the readability of the content displayed on the interface, and the adaptability of the software across different scenarios and conditions.

The selection of tasks was not done in isolation but was aligned with industry standards such as IEC 62366. This alignment ensured that the tasks were not only relevant but also met the expectations and requirements of the industry. The tasks were carefully crafted to simulate real-world usage scenarios. This simulation was crucial to ensure a thorough examination of the SaMD’s usability in diverse situations that users might encounter in their daily interactions with the software.

The tasks selected for the usability study are detailed below. Each task is accompanied by its respective objectives. This combination of tasks and objectives provides a robust framework for the subsequent evaluation of the software’s performance and user experience. This framework is essential to ensure that the evaluation is not only comprehensive but also objective and based on measurable criteria.

### 4.1 Healthcare Personnel

During the usability study, healthcare professionals were tasked with a series of activities to assess the Software as a Medical Device (SaMD). These activities were not random but were carefully chosen to cover a wide range of functions and features of the software. The activities included creating and sending questionnaire links to patients, interpreting patient details within the platform, sending multiple Patient-Reported Outcome Measures (PROMs) to the same patient, checking updated patient data, confirming device compatibility, performing common login and task sequences, finding and utilising support resources, updating and saving patient data, and sending reminders to patients.

The objectives of these activities ranged from evaluating ease of use and efficiency to ensuring compatibility and discoverability of support features. These objectives were not only diverse but also comprehensive, covering a wide range of aspects related to the software’s usability. The evaluation of ease of use and efficiency, for instance, was crucial to ensure that healthcare professionals could use the software without unnecessary difficulties or delays. The assessment of compatibility and discoverability of support features, on the other hand, was essential to ensure that the software could be used on different devices and that users could easily find and utilise the support resources provided.

**Table 2.**
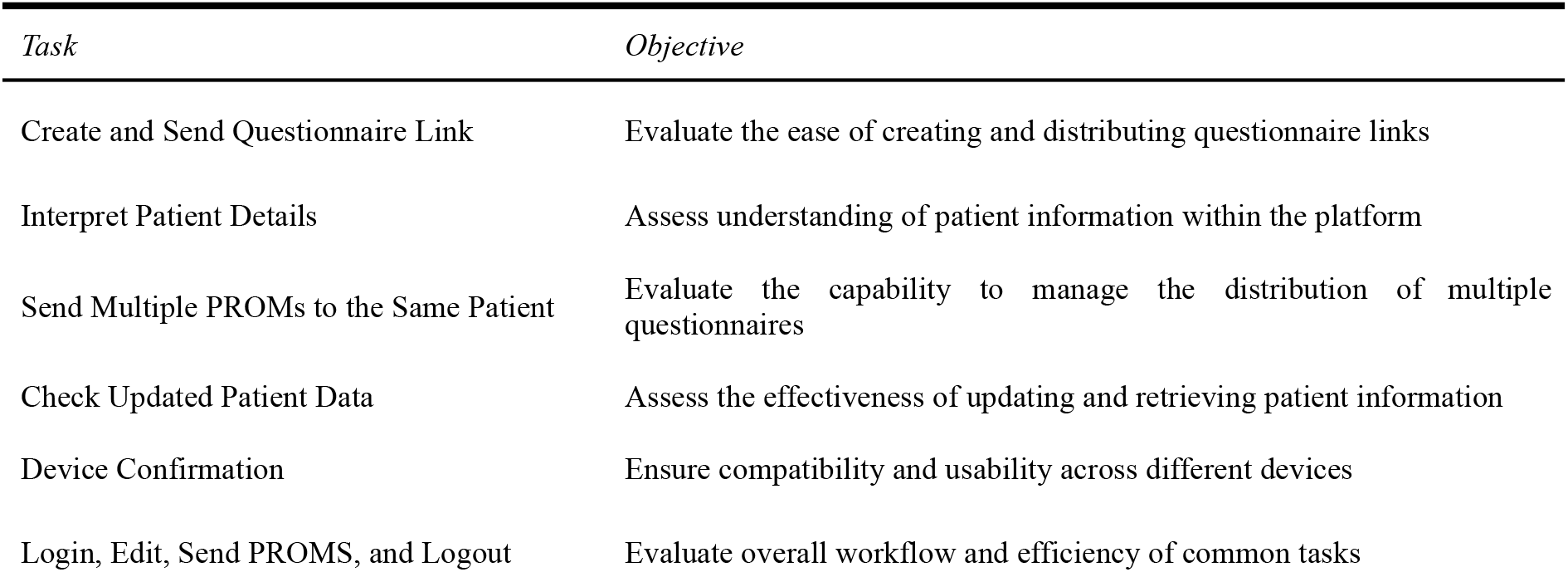
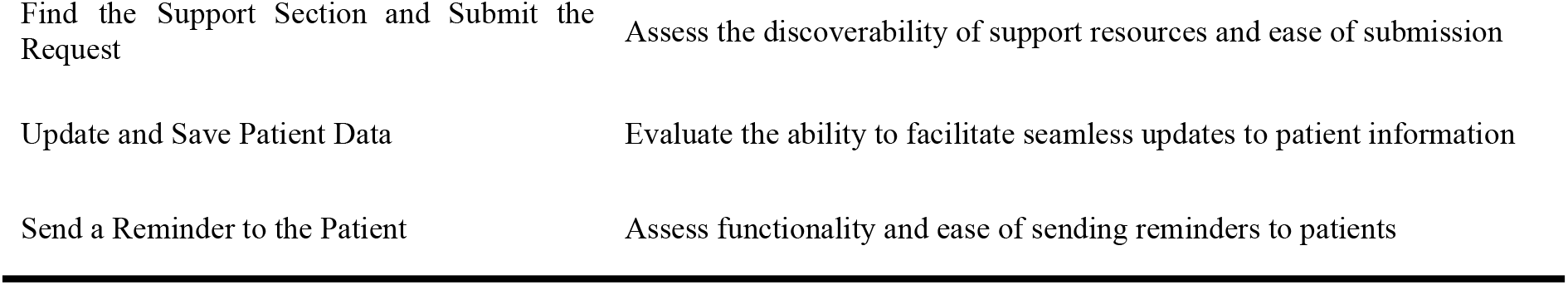
User Group: Healthcare Personnel.

| Table 2 |  | User Group: Healthcare Personnel. |
| --- | --- | --- |
| Task | Objective |  |
| Create and Send Questionnaire Link | Evaluate the ease of creating and distributing questionnaire links |  |
| Interpret Patient Details | Assess understanding of patient information within the platform |  |
| Send Multiple PROMs to the Same Patient | Evaluate the capability to manage the distribution of multiple questionnaires |  |
| Check Updated Patient Data | Assess the effectiveness of updating and retrieving patient information |  |
| Device Confirmation | Ensure compatibility and usability across different devices |  |
| Login, Edit, Send PROMS, and Logout | Evaluate overall workflow and efficiency of common tasks |  |

|  |  |
| --- | --- |
| Find the Support Section and Submit the Request | Assess the discoverability of support resources and ease of submission |
| Update and Save Patient Data | Evaluate the ability to facilitate seamless updates to patient information |
| Send a Reminder to the Patient | Assess functionality and ease of sending reminders to patients |

### 4.2 General Population

During the usability study, participants from the general population were assigned a series of tasks to assess the software’s performance. These tasks were not random but were carefully chosen to cover a wide range of functions and features of the software. The tasks included checking the software’s usability in both dark and light modes, reading content provided through links, answering a questionnaire and submitting responses, hovering over and clicking on designated areas to evaluate interface responsiveness, rating the perceived difficulty of answering questions, and filling out the questionnaire at different screen resolutions.

The objectives of these tasks ranged from evaluating visual preferences and readability to assessing user interaction precision and adaptability to various devices. These objectives were not only diverse but also comprehensive, covering a wide range of aspects related to the software’s usability. The evaluation of visual preferences and readability, for instance, was crucial to ensure that users could easily read and understand the content displayed on the interface. The assessment of user interaction precision and adaptability to various devices, on the other hand, was essential to ensure that users could accurately interact with the interface and that the software could adapt to different devices and screen resolutions.

**Table 3.**
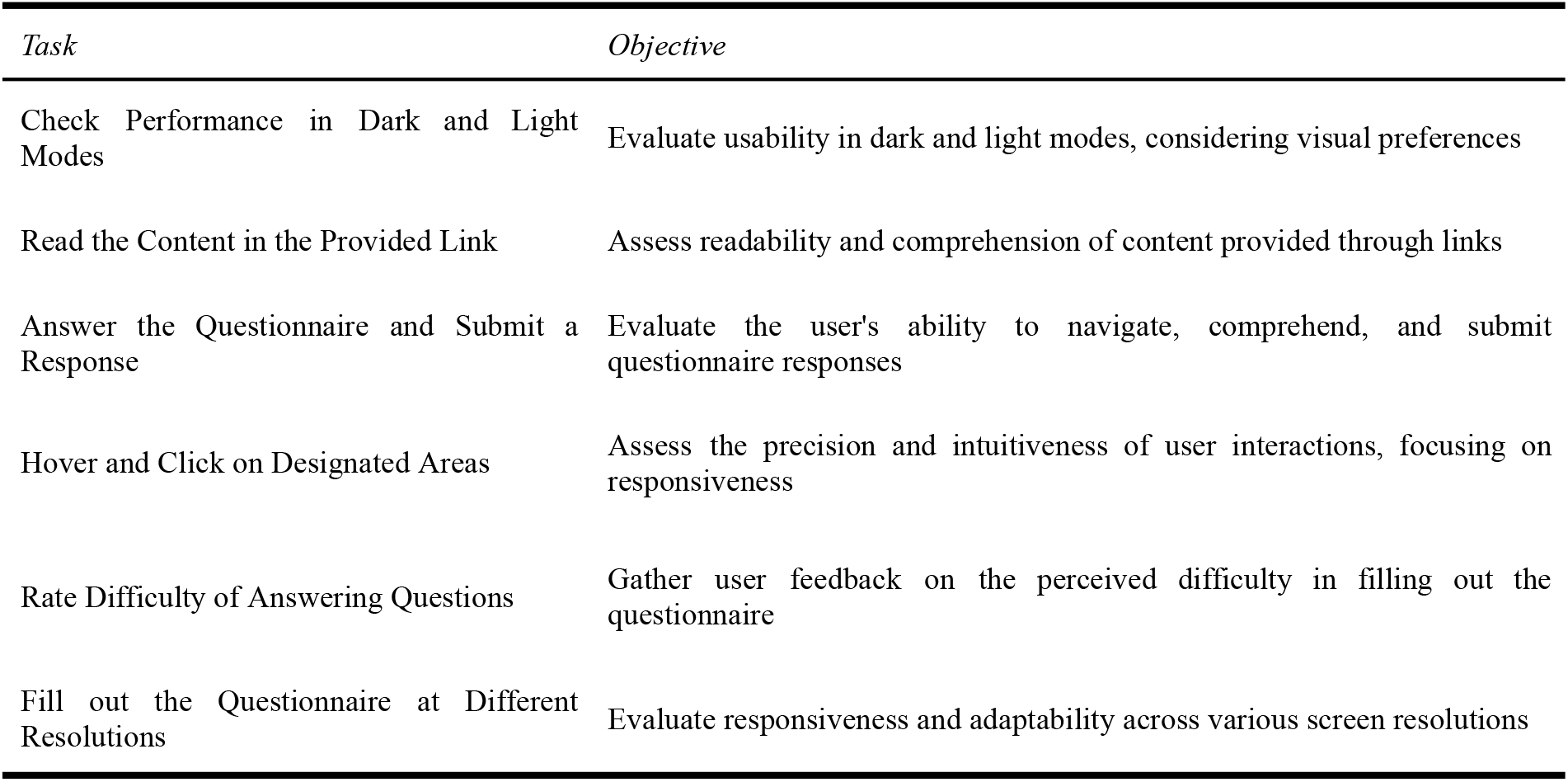
User Group: General Public.

## 5 Task Evaluation

The tasks selected for the evaluation phase were meticulously designed to cover critical functionalities, ensuring a thorough examination of the Software as a Medical Device’s (SaMD) usability. These tasks were not chosen randomly but were carefully crafted to simulate real-world scenarios that potential software users might encounter. This was done to ensure that the evaluation was as comprehensive and realistic as possible and that it would provide a true reflection of the software’s performance in a variety of situations.

The tasks incorporated elements such as dark and light modes, link navigation, questionnaire completion, and interface responsiveness across various resolutions. These elements were chosen because they are representative of the types of functionalities that users might need to use in a real-world setting. For example, the ability to switch between dark and light modes is a feature that many users find useful, especially those who use software for extended periods of time. Similarly, the ability to navigate through links and complete questionnaires are functionalities that are often required in healthcare settings.

Both healthcare professionals and the general population participated in the study, providing a diverse and inclusive representation of potential end-users. This was done to ensure that the evaluation was not biased towards any particular group of users and that it would provide a true reflection of the software’s performance across a wide range of user types. Including both healthcare professionals and the general population in the study also ensured that the evaluation would consider the different needs and requirements of these two groups.

The task evaluation results were notably positive, with a 100% success rate observed for both user groups. This outcome underscores the high usability of the SaMD, as reflected in the successful completion of all assigned tasks. The fact that all participants were able to complete all tasks successfully indicates that the software is user-friendly and easy to use, even for those who may not be familiar with this type of technology.

Detailed metrics were recorded during the evaluation, capturing counts of successful and unsuccessful test instances. These metrics provided a clear and objective measure of the software’s performance, allowing for a thorough and accurate assessment of its usability. Notably, no use errors or hazards were identified during the evaluation, affirming the software’s reliability and robust design.

Participants demonstrated proficiency in tasks such as checking performance in dark and light modes, reading content through provided links, completing questionnaires, and navigating the interface through hovering and clicking actions. The software exhibited consistent and reliable performance across various screen resolutions, further validating its adaptability to diverse user environments.

The positive task evaluation outcomes provide strong evidence of the software’s effectiveness and user-friendliness. These results align with industry standards and contribute valuable insights into the SaMD’s performance, paving the way for confident assertions regarding its usability and reliability in real-world healthcare settings. The fact that the software performed well in all tasks and that no use errors or hazards were identified provides strong evidence of its reliability and robustness. This, combined with the high success rate observed for both user groups, indicates that the software is effective, user-friendly, and easy to use.

## 6 Software Overview

The Pathpoint Outcomes Software, a groundbreaking technological advancement in the healthcare industry, is a product of Open Medical, a leading provider of cutting-edge healthcare technology. This innovative software solution operates on a cloud-based, Software-as-a-service (SaaS) model. This model offers significant advantages in terms of scalability, accessibility, and cost-effectiveness, making it a preferred choice for healthcare providers looking to enhance their service delivery.

The cloud-based, Software-as-a-service (SaaS) model that the Pathpoint Outcomes Software operates on is a revolutionary approach in the healthcare industry. This model allows for the software to be accessed from any location and at any time, providing healthcare providers with the flexibility they need to deliver high-quality care. In addition, the SaaS model offers scalability, allowing healthcare providers to easily expand or reduce their usage based on their needs. This scalability ensures that healthcare providers only pay for what they use, making the software a cost-effective solution.

Another significant advantage of the SaaS model is its accessibility. With the software being cloud-based, healthcare providers can access it from any device with an internet connection. This accessibility ensures that healthcare providers can access patient information quickly and easily, enhancing the quality of care provided. Moreover, the SaaS model eliminates the need for healthcare providers to invest in expensive hardware or software, further enhancing its cost-effectiveness.

A key feature of the Pathpoint Outcomes Software is its seamless integration with electronic healthcare records. This integration ensures smooth and efficient data exchange, eliminating the potential for errors that can occur with manual data entry. By automating data exchange, the software reduces the time and effort required by healthcare providers to access and update patient information. This efficiency not only enhances the quality of care provided but also allows healthcare providers to focus more on patient care rather than administrative tasks.

The seamless integration of the Pathpoint Outcomes Software with electronic healthcare records also ensures that patient information is up-to-date and accurate. This accuracy is crucial in healthcare, as it ensures that healthcare providers have the most recent and accurate information when making decisions about patient care. By ensuring the accuracy of patient information, the software enhances the effectiveness of healthcare delivery, leading to improved patient outcomes.

Regarding security, the Pathpoint Outcomes Software aligns with industry-standard security protocols. This alignment ensures that sensitive patient data is protected from potential threats, demonstrating Open Medical’s commitment to data privacy and security. By aligning with industry-standard security protocols, the software ensures that patient data is stored and transmitted securely, reducing the risk of data breaches.

The software’s compliance with NHS Digital standards further validates its reliability and effectiveness. These standards set out stringent requirements for healthcare technology, covering aspects such as interoperability, data quality, and security. By meeting these requirements, the Pathpoint Outcomes Software demonstrates its ability to deliver reliable, high-quality service. The compliance with NHS Digital standards also ensures that the software is compatible with other healthcare technologies, enhancing its interoperability.

In addition to complying with NHS Digital standards, the software adheres to the Web Content Accessibility Guidelines. These guidelines ensure the software is accessible to all users, including those with disabilities. This commitment to accessibility underscores Open Medical’s dedication to making healthcare more inclusive. By adhering to the Web Content Accessibility Guidelines, the software ensures that all users, regardless of their abilities, can access and use the software effectively.

Finally, the software has received high ratings for the DTAC criteria by NHS England and ORCHA, a prominent body assessing healthcare technology’s quality and effectiveness. These high ratings provide further testament to the software’s high performance and reliability. The high ratings from NHS England and ORCHA also demonstrate the software’s effectiveness in improving healthcare delivery, further validating its value to healthcare providers.

In summary, the Pathpoint Outcomes Software is a robust, reliable, and effective solution for healthcare providers. Its seamless integration with electronic healthcare records, compliance with stringent industry standards, and commitment to accessibility and security make it an excellent choice for healthcare providers looking to enhance their service delivery. With its innovative features and proven effectiveness, the Pathpoint Outcomes Software is set to revolutionize healthcare delivery, leading to improved patient outcomes and enhanced healthcare efficiency.

## 7 Results of Quantitative and Qualitative Outcomes

The study’s results indicated a strong positive outcome for the software’s usability, both in terms of quantitative and qualitative data. The quantitative data was particularly compelling, with a usability rating of 95% obtained from the Digital Therapeutics Assessment and Certification (DTAC). This high rating is a testament to the software’s user-friendly nature, indicating that users found it easy to navigate and interact with. The DTAC is a well-respected organisation that assesses and certifies digital therapeutics, so their high rating carries significant weight and credibility.

In addition to the DTAC’s rating, the Organisation for the Review of Care and Health Applications (ORCHA) also gave the software a high usability rating of 85%. ORCHA is another reputable organisation that reviews and rates health applications, so their rating further validates the software’s user-friendly nature. The fact that two separate organisations both gave the software high usability ratings is a strong indication of its user-friendliness and ease of use.

The study also revealed a 100% success rate in using the software. This means that all users were able to successfully navigate and utilise the software without encountering any major issues or obstacles. This success rate aligns with the high usability ratings obtained from DTAC and ORCHA, further reinforcing the software’s user-friendly nature. It is clear from this data that the software is not only easy to use, but also effective in achieving its intended purpose.

The qualitative feedback obtained from Healthcare Professionals and the General Population also emphasised the software’s intuitiveness. Healthcare Professionals are typically highly trained and experienced in using a variety of software and technology in their field. Their feedback is therefore particularly valuable, as they have a high level of expertise and understanding of what makes a software user-friendly and effective. They praised the software for its ease of use and intuitive design, indicating that it was easy to incorporate into their daily workflow.

The General Population’s feedback was similarly positive, highlighting the software’s intuitiveness and user-friendly interface. These users found the software easy to navigate and understand, even without any prior knowledge or experience using similar software. This feedback suggests that the software is accessible to a wide range of users, not just those with technical expertise. It is clear from this feedback that the software’s design is intuitive and user-friendly, making it easy for users of all levels of expertise to navigate and use.

Despite the overwhelmingly positive feedback, there were also suggestions for further improvements. Both Healthcare Professionals and the General Population provided feedback on areas where the software could be enhanced to better meet their needs. This feedback is incredibly valuable, as it provides insights into how the software could be improved in future updates and iterations. By taking this feedback into account, the software can continue to evolve and improve, ensuring that it remains user-friendly and effective.

In summary, the study demonstrated that the software had a high usability rating, with quantitative data indicating a 95% usability rating on DTAC and an 85% rating by ORCHA. The 100% success rate in using the software further confirmed its user-friendliness. The qualitative feedback from Healthcare Professionals and the General Population emphasised the software’s intuitiveness and provided valuable insights for further improvements. The study’s results are a testament to the software’s user-friendly nature and effectiveness, and provide a strong foundation for future improvements and iterations.

## 8 Conclusion

The Usability Evaluation is a crucial tool that affirms the usability characteristics of the Pathpoint Outcomes Software, a leading-edge platform in the healthcare sector. This evaluation is not just a cursory review; it is a comprehensive and in-depth assessment that guides ongoing enhancements to ensure the software is safe, effective, and user-friendly. The primary objective of this evaluation is to provide a thorough understanding of the software’s usability, thereby facilitating improvements that will enhance its performance and user experience.

This evaluation focuses on various usability characteristics of the Pathpoint Outcomes Software. It delves into the intricate details of the software, examining it from multiple perspectives to ensure it is efficient, effective, and user-friendly. This is achieved by scrutinizing the software’s features, design, and functionality, and by considering how different types of users, each with their unique needs and preferences, interact with it. This multi-faceted approach to evaluation provides a holistic view of the software’s usability, offering a more accurate and comprehensive understanding of its strengths and weaknesses.

The primary goal of this evaluation is to guide ongoing enhancements in the Pathpoint Outcomes Software. By identifying areas where the software excels and areas that need improvement, the evaluation plays a pivotal role in shaping the future development of the software. This process of continuous improvement, driven by the insights gained from the evaluation, ensures that the software remains safe, user-friendly, and effective. It also ensures that the software continues to meet the evolving needs of its users, thereby enhancing its value and relevance in the healthcare sector.

The Pathpoint Outcomes Software is designed for use by a diverse range of user groups, including healthcare professionals, administrators, and patients. This diversity is a key consideration during the evaluation process. The evaluation ensures that the software is not only accessible to all potential users but also that it is useful and intuitive, regardless of the user’s technical expertise or familiarity with healthcare software. The results of the study offer significant insights into the rapidly evolving landscape of healthcare software usability. By understanding how the software is used in real-world situations, developers can continue to refine and improve the software, enhancing the user experience and ensuring the software remains relevant and effective in the dynamic healthcare environment.

In addition to informing the development of the Pathpoint Outcomes Software, the insights gained from the usability evaluation are also valuable for the broader field of healthcare software. They provide a window into the current state of usability in the sector, offering lessons that can be applied to other software solutions. This not only helps improve the Pathpoint Outcomes Software but also contributes to the ongoing evolution and improvement of healthcare software in general. By sharing these insights, the evaluation plays a crucial role in driving innovation and improvement in the healthcare software sector, benefiting not just the users of the Pathpoint Outcomes Software but also the wider healthcare community.

In summary, the Usability Evaluation is a vital tool that ensures the Pathpoint Outcomes Software is safe, effective, and user-friendly. It guides ongoing enhancements, considers the needs of diverse user groups, and provides valuable insights into the evolving landscape of healthcare software usability. The insights gained from this evaluation are not only beneficial for the development of the Pathpoint Outcomes Software but also contribute to the broader field of healthcare software.

## Supporting information

Usability questionnaire and Hazard list

Risk file

## Data Availability

The data is available on reasonable request

## Acknowledgements

We would like to take this opportunity to express our profound gratitude to all the participants who took part in our Software as a Medical Device (SaMD) usability study. Their valuable contributions, insightful feedback, and active participation played a pivotal role in the success of our research. We are deeply appreciative of their time, effort, and commitment to the study. Special thanks are due to Open Medical Ltd for their unwavering support and constant encouragement throughout the research process. We would like to particularly acknowledge Mr. Jed Aubrey, whose guidance and expertise significantly enhanced the overall quality and success of the study. His dedication and commitment to our research were truly remarkable. We are immensely grateful for their support, which was instrumental in the successful completion of our study.

