## Supplementary material for "Comprehensive Usability Evaluation and Performance Overview of Pathpoint® Outcomes Software": Usability questionnaire and Hazard list: OM_Medical Device_List of Hazard-Related Use Scenarios.pdf

OpenMedical

CP House, 97 - 107 Uxbridge Road, London, W5 5TL, United Kingdom
www.openmedical.co.uk   +44 (0)20 3475 2955

### List of Hazard-Related Use Scenarios

RESTRICTED

CP House, 97 - 107 Uxbridge Road, London, W5 5TL, United Kingdom
www.openmedical.co.uk +44 (0)20 3475 2955

#### Document Control

| | |
|---|---|
| Document Title | OM_Medical Device_List of Hazard-Related Use Scenarios |
| Written By | Atlas Naqvi and Muna Mohamud |
| Reviewed By | Balamurugan Subramaniyan |
| Approved By | Piyush Mahapatra |
| Published Date | 14-Dec-2023 |
| Effective Date | 14-Dec-2023 |
| Review Cycle / Date | Annually |
| Document Classification | RESTRICTED |
| Distribution | Executive Team |

##### Document History

| Issue | Date | Author | Change Log |
|---|---|---|---|
| V1.0 | 14-Dec-2023 | Balamurugan Subramaniyan | Initial Release |

### Contents

Summary 4
Mapping of Standard Requirements to Document Sections 4
Hazard-Related Use Scenarios 4

### Summary

The goal of the list of Hazard-Related Use Scenarios is to gather all Use Scenarios that could be associated with a Hazard, e.g. if a user doesn't understand a particular feature of the Pathpoint Outcome software. A Hazard could be encountered, subsequently leading to a Hazardous Situation.

#### Mapping of Standard Requirements to Document Sections

| IEC 62366-1:2015 Section | Title | Document Section |
|---|---|---|
| 5.4 | Identify and describe Hazard-Related Use Scenarios | 1 |
| 5.5 | Select the Hazard-Related Use Scenarios for Summative Evaluation | 1 |

### Hazard-Related Use Scenarios

By the principles outlined in IEC 62366, our hazard-related use scenario analysis is designed to comprehensively address the potential risks of using our medical device within two distinct user populations: healthcare professionals (HCPs) and the broader general public. For the HCP group, we focus on understanding the specific tasks, interactions, and potential hazards that may arise in a clinical or professional setting. This includes considerations for healthcare professionals' unique expertise, training, and environment. Simultaneously, we diligently evaluate hazard-related use scenarios for the general public to identify and mitigate risks that may emerge in non-professional settings. Our goal is to develop a user-centered approach that addresses the diverse needs and capabilities of both user groups, ensuring the safety and effectiveness of our medical device across various usage contexts.

**Table 1: User Group: Healthcare Personnel.**

| ID | Description of risk | Tasks | Acceptance Criteria |
|---|---|---|---|
| 1 | End-to-end usage from login to sign-out | Ask the HCP to perform end-to-end task | The HCP should be able to perform the tasks comfortably |
| 2 | Visual difficulty in the monitor may impede healthcare professionals' ability to interpret and respond to critical patient information accurately | Ask the HCP to interpret the details of the patient that is present in the platform | The HCP should be able to interpret the details of the patients correctly |
| 3 | The HCP does not know how to save the data of the patient after updating | Ask the HCP to update a patient's data and keep it | The HCP should be able to edit patient's details and save them |

| | | | |
|---|---|---|---|
| 4 | The HCP sent the Questionnaire link to the wrong person | Ask the HCP to send the questionnaire link to the patient of choice | The HCP should be able to send the link to the correct patient |
| 5 | The HCP is not able to send multiple PROMS questionnaires to the same patient | Ask the HCP to create the questionnaire link that contains multiple PROMS | The HCP should be able to incorporate multiple PROMS and be able to send the link to the right user |
| 6 | The HCP does not know how to send a PROMS reminder to the patients | Ask the HCP to send the reminder to the patient to fill in their PROMS questionnaire | The HCP should be able to send a reminder to the patients |
| 7 | The HCP is not able to see the update after the patient has successfully filled in the PROMS questionnaire | Ask the HCP if they can check the patient data after updating the information | The HCP should be able to see the progress after the PROMS are answered |
| 8 | The HCP does not know how to contact the Open Medical Support team from the platform | Ask the HCP about where to find the support section in the software | The HCP should be able to click on the support button on the platform |
| 9 | HCP is not able to use the software platform with consistency between device variants (iPad, laptop) | Ask different HCPs to use the platform on more than one type of device and compare the consistency of usage | The HCP has to be satisfied with the usage of the platform on different devices |

**Table 2: User Group: General Public**

| ID | Description | Tasks | Acceptance Criteria |
|---|---|---|---|
| 1 | The device does not perform the same way in dark and light mode | Ask about the performance of the software in different modes | The patient should be able to use the software comfortably without any difficulty |
| 2 | Visual difficulty in the monitor may impede the patient from accurately interpreting and responding to critical things in a questionnaire | Ask the patient if they had any visual difficulty accessing the software | The patient should not complain of any visual difficulty while using the software |

CP House, 97 - 107 Uxbridge Road, London, W5 5TL, United Kingdom
www.openmedical.co.uk +44 (0)20 3475 2955

| | | | |
|---|---|---|---|
| 3 | The patient filled in the answers to the questionnaire and does not know how to submit their responses | Check if the patient can submit the response after answering all the questions | The patient should be able to submit the response after filling out the questionnaire |
| 5 | Patient discomfort using their device during the response submission process may hinder their ability to provide accurate and timely information. | Ask the patient how comfortable they are when using their device to respond to the questions | The patient should not feel any discomfort in using their device and scrolling |
| 7 | The resolution of the questionnaire screen in different formats of response (SMS, email) | Check how different patients felt about different resolutions | The patient should not complain of any visual difficulty while using the software |
| 8 | The patient can complete multiple PROMS with ease | Check if the patient has received all PROMS sent and can complete them | The patient should be able to access all PROMS and complete them with ease |
