## Supplementary material for "Comprehensive Usability Evaluation and Performance Overview of Pathpoint® Outcomes Software": Usability questionnaire and Hazard list: OM_Medical Device_Usability Questionnaire General Population.pdf

CP House, 97 - 107 Uxbridge Road, London, W5 5TL, United Kingdom
www.openmedical.co.uk +44 (0)20 3475 2955

### Usability Questionnaire General Population

CP House, 97 - 107 Uxbridge Road, London, W5 5TL, United Kingdom
www.openmedical.co.uk +44 (0)20 3475 2955

#### Document Control

| | |
|---|---|
| Document Title | OM_Medical Device_Usability Questionnaire General Population |
| Written By | Balamurugan Subramaniyan |
| Approved By | Atlas Naqvi and Muna Mohamud |
| Published Date | 14-Dec-2023 |
| Effective Date | 14-Dec-2023 |
| Review Cycle / Date | |
| Document Classification | RESTRICTED |
| Distribution | Executive Team |

##### Document History

| Issue | Date | Author | Change Log |
|---|---|---|---|
| V1.0 | 14-Dec-2023 | Balamurugan Subramaniyan | Initial Release |

### Contents

Usability Questionnaire General Population 1
1 General Information (To be filled by Moderator) 4
2 Instruction for User 4
3 Quantitative Section (Rate from 1 – 5) 4
4 Qualitative Section (Please add any description in the comments section) 6
5 Moderator's Comment/Observation 7

### 1 General Information (To be filled by Moderator)

| | |
|---|---|
| Name of the User | |
| Phone Number | |
| Address or Name of Hospital | |
| Date | MM-DD-YYYY |
| Is the user trained? | Yes or No |
| Name of the Device | |
| Version number of software/app | |
| Name of Moderator | |

### 2 Instruction for User

- Complete this questionnaire once you have familiarized yourself with the device and software application
- Ensure you have read the user manual
- Ensure you answer all the questions independently and to the best of your knowledge
- In case you may have any questions and seek clarity please contact the moderator present during the study
- Please provide complete details in the answers you provide

In case of any doubt, please ask the moderator before answering any question

### 3 Quantitative Section (Rate from 1 – 5)

3.1 Rate the clarity and intuitiveness of the user interface elements (e.g., buttons, menus).
(1 = Not at all comfortable and 5 = Very comfortable)

- ☐ 1
- ☐ 2
- ☐ 3
- ☐ 4
- ☐ 5

3.2 How would you rate the system's responsiveness during the questionnaire completion?
(1 = Not at all comfortable and 5 = Very comfortable)

- ☐ 1
- ☐ 2
- ☐ 3
- ☐ 4

- ☐ 5

3.3 Rate the clarity and user-friendliness of the interface provided for accessing and responding to the questionnaire. (1 = Very Difficult and 5 = Very Easy)

- ☐ 1
- ☐ 2
- ☐ 3
- ☐ 4
- ☐ 5

3.4 Did you experience any delays or lag while interacting with the system?
(1 = Very Difficult and 5 = Very Easy)

- ☐ 1
- ☐ 2
- ☐ 3
- ☐ 4
- ☐ 5

3.5 How quickly did you understand how to access and complete the questionnaire?
(1 = Very Difficult and 5 = Very Easy)

- ☐ 1
- ☐ 2
- ☐ 3
- ☐ 4
- ☐ 5

3.6 How confident are you that your responses were accurately recorded?
(1 = Very Difficult and 5 = Very Easy)

- ☐ 1
- ☐ 2
- ☐ 3
- ☐ 4
- ☐ 5

3.7 How would you rate the overall usability of the system for your daily tasks?

CP House, 97 - 107 Uxbridge Road, London, W5 5TL, United Kingdom
www.openmedical.co.uk +44 (0)20 3475 2955

(1 = Very Difficult and 5 = Very Easy)

- ☐ 1
- ☐ 2
- ☐ 3
- ☐ 4
- ☐ 5

### 4 Qualitative Section (Please add any description in the comments section)

4.1 Can you provide feedback on your overall experience completing the questionnaire?

4.2 How do you feel about the system's role in empowering you to actively participate in your healthcare by responding to questionnaires?

### 5 Moderator's Comment/Observation

The Moderator should list his or her observations based on his/her interaction with the user during this session.

Signature of the User
Date: MM-DD-YYYY

Signature of Moderator
Date: MM-DD-YYYY
