## Supplementary material for "Comprehensive Usability Evaluation and Performance Overview of Pathpoint® Outcomes Software": Usability questionnaire and Hazard list: OM_Medical Device_Usability Questionnaire HCP.pdf

CP House, 97 - 107 Uxbridge Road, London, W5 5TL, United Kingdom
www.openmedical.co.uk +44 (0)20 3475 2955

### Usability Questionnaire HCP

#### Document Control

RESTRICTED

| | |
|---|---|
| Document Title | OM_Medical Device_Usability Questionnaire HCP |
| Written By | Atlas Naqvi and Muna Mohamud |
| Reviewed By | Balamurugan Subramaniyan |
| Approved By | Piyush Mahapatra |
| Published Date | 16-Dec-2023 |
| Effective Date | 16-Dec-2023 |
| Review Cycle / Date | Annually |
| Document Classification | RESTRICTED |
| Distribution | Executive Team |

Document History

| Issue | Date | Author | Change Log |
|---|---|---|---|
| V1.0 | 16-Dec-2023 | Balamurugan Subramaniyan | Initial Release |

CP House, 97 - 107 Uxbridge Road, London, W5 5TL, United Kingdom
www.openmedical.co.uk +44 (0)20 3475 2955

### Contents

Usability Questionnaire HCP 1
1 General Information (To be filled by Moderator) 4
2 Instruction for User 4
3 Quantitative Section (Rate from 1 – 5) 4
4 Qualitative Section (Please add any description in the comments section) 6
5 Moderator's Comment/Observation 7

### 1 General Information (To be filled by Moderator)

| | |
|---|---|
| Name of the User | |
| Phone Number | |
| Occupation | |
| Location | |
| Date | MM-DD-YYYY |
| Is the user trained? | Yes or No |
| Name of the Device | |
| Version number of software/app | |
| Name of Moderator | |

### 2 Instruction for User

- Complete this questionnaire once you have familiarised yourself with the device and software application
- Ensure you have read the user manual
- Ensure you answer all the questions independently and to the best of your knowledge
- In case you may have any questions and seek clarity, please contact the moderator present during the study
- Please provide complete details in the answers you provide

In case of any doubt, please ask the moderator before answering any question

### 3 Quantitative Section (Rate from 1 – 5)

3.1 Rate the clarity and intuitiveness of the user interface elements (e.g., buttons, menus).
(1 = Not at all comfortable and 5 = Very comfortable)

- ☐ 1
- ☐ 2
- ☐ 3
- ☐ 4
- ☐ 5

3.2 How would you rate the efficiency of retrieving patient information from the system?
(1 = Not at all comfortable and 5 = Very comfortable)

- ☐ 1
- ☐ 2
- ☐ 3
- ☐ 4

☐ 5

3.3 Rate the ease with which you could navigate between different system sections.
(1 = Very Difficult and 5 = Very Easy)

☐ 1

☐ 2

☐ 3

☐ 4

☐ 5

3.4 How easily could you recover from errors or incorrect entries within the system?
(1 = Very Difficult and 5 = Very Easy)

☐ 1

☐ 2

☐ 3

☐ 4

☐ 5

3.5 Did you experience any delays or lag while interacting with the system?
(1 = Very Difficult and 5 = Very Easy)

☐ 1

☐ 2

☐ 3

☐ 4

☐ 5

3.6 How easy is it to understand the user manual?
(1 = Very Difficult and 5 = Very Easy)

☐ 1

☐ 2

☐ 3

☐ 4

☐ 5

3.7 How would you rate the ease and speed with which you could complete everyday tasks (e.g., retrieving patient information, entering clinical notes)?

(1 = Very Difficult and 5 = Very Easy)

- ☐ 1
- ☐ 2
- ☐ 3
- ☐ 4
- ☐ 5

3.8 How would you rate the system's overall usability for your daily tasks?
(1 = Very Difficult and 5 = Very Easy)

- ☐ 1
- ☐ 2
- ☐ 3
- ☐ 4
- ☐ 5

### 4 Qualitative Section (Please add any description in the comments section)

4.1 Can you provide your feedback on the ease of integrating multiple PROMS and sending it to the user?

4.2 Please provide your feedback on your end-to-end usage of the platform.

CP House, 97 - 107 Uxbridge Road, London, W5 5TL, United Kingdom
www.openmedical.co.uk +44 (0)20 3475 2955

### 5 Moderator's Comment/Observation

The Moderator should list his or her observations based on his/her interaction with the user during this session.

Signature of the User
Date: MM-DD-YYYY

Signature of Moderator
Date: MM-DD-YYYY
